# The Accuracy of Quantitative Lutetium-177 Single Photon Emission Computed Tomography Imaging and the Role for Partial-Volume Corrections in Clinical Practice: A Systematic Review

**DOI:** 10.1101/2025.06.09.25329310

**Authors:** Julian C. J. van Oorschodt, Guy D. Eslick, Martin Ugander, Carola van Pul, Enid M. Eslick

**Affiliations:** Faculty of Medicine and Health, The University of Sydney, Sydney, New South Wales, Australia; Department of Applied Physics, Eindhoven University of Technology, Eindhoven, The Netherlands; Clinical Links Using Evidence-based Data (CLUED) Consulting Pty Ltd, Sydney, New South Wales, Australia

**Keywords:** Lutetium, partial-volume corrections, partial-volume effects, peptide receptor radionuclide therapy, quantitative imaging, SPECT/CT

## Abstract

**Background:** Accurate quantification of lutetium-177 (^177^Lu) radioactivity in SPECT/CT imaging is essential for a further development of ^177^Lu-based peptide receptor radionuclide therapy (PRRT) for neuroendocrine tumours and metastatic prostate cancer.

**Purpose:** This review provides an overview on the accuracy of quantitative ^177^Lu SPECT/CT imaging, identifies methods for an improved accuracy when imaging small volumes subject to partial-volume effects (PVEs), and assesses the role for partial-volume corrections (PVCs) in clinical practice.

**Methods:** A systematic review was conducted according to PRISMA guidelines. MEDLINE, EMBASE, and Web of Science databases were searched with no language restrictions. Original studies explicitly reporting on the accuracy of ^177^Lu activity quantifications in physical SPECT/CT measurements were included.

**Results:** The literature search identified 616 records, of which 46 studies were included for analysis. Percentage errors of quantifications were found to constitute a large range (−102% to 285%). The recovery of ^177^Lu activity from small volumes is inherently limited by PVEs. The application of PVCs has led to improvements in the accuracy and precision of quantifications on small volumes in phantom imaging.

**Conclusion:** The accuracy of ^177^Lu activity quantifications in SPECT/CT imaging is subject to large variability and will be degraded by PVEs when imaging small volumes. The data suggests that the implementation of standardised procedures and PVCs may lead to an improved accuracy and precision of quantitative ^177^Lu SPECT/CT imaging in clinical practice, thereby allowing the further development of ^177^Lu-based PRRT.

## I Introduction

Lutetium-177 (^177^Lu)-based peptide receptor radionuclide therapy (PRRT) is a promising therapy for neuroendocrine tumours and metastatic castration-resistant prostate cancer.^1–3^ Further developments of the therapy include the implementation of individualised treatment plans^4–6^, and will require an improved understanding of tumour dose - treatment response relationships.^7–9^ Accurate quantifications of ^177^Lu radioactivity in SPECT/CT images will be crucial for the reliable determination of this relationship through large scale dosimetry analyses.^10–13^

^177^Lu is an attractive radionuclide used in PRRT due to its favourable physical properties that allow for both effective treatment and efficient imaging of tumours.^14–16^ Through *β*^−^-emission with a short mean path length of 0.7 mm, ^177^Lu can deliver high doses of radiation to tumours while limiting damage to neighbouring healthy tissues. Additionally, with *γ*-rays predominantly being emitted at energies of 113 and 208 keV with abundances of 6.2 and 10.4%, respectively, ^177^Lu is also suitable for direct visualisation via commercial SPECT systems.

Single-photon emission computed tomography (SPECT) imaging, unlike positron emission tomography (PET) imaging, which has been designed to be quantitative, has long been considered a purely qualitative modality.^17–19^ In recent decades however, various technological advances have enabled the acquisition of quantitative data through SPECT imaging.^10, 14, 20^ With the integration of a CT component intothe SPECT modality to create the SPECT/CT system, sophisticated correction methods have been incorporated into reconstruction algorithms to address processes such as photon attenuation and scatter. These developments have also made it possible to derive a conversion factor that transforms a reconstructed SPECT/CT image from counts per pixel into units of activity concentration per unit volume (Bq/mL).^18^

Nevertheless, quantitative ^177^Lu SPECT/CT imaging is not yet widely established, partly because quantification remains a time-intensive process.^5, 6, 15^ Additionally, the absence of a widely accepted standardized procedure for quantitative ^177^Lu SPECT/CT imaging has resulted in inconsistencies across imaging hardware, calibration methods, acquisition protocols, and reconstruction algorithms.^9, 21, 22^ As a result, this has led to variability in the accuracy of quantitative ^177^Lu SPECT/CT imaging ^7, 14, 16, 20^, hindering the ability to compare quantitative data across multicenter studies.^23^

Ideally, standardised protocols for quantitative ^177^Lu SPECT/CT imaging in clinical practice will include partial-volume corrections (PVCs) to account for displacements of counts between different image regions due to partial-volume effects (PVEs).^11^ In SPECT/CT imaging, PVEs originate from a limited (i.e., in the centimetre range) spatial resolution of commercial SPECT/CT systems, and affect quantifications on objects smaller than approximately 3 times the spatial resolution of the system.^17^ Consequently, PVEs constitute one of the largest sources of inaccuracy in the clinical practice of quantitative ^177^Lu SPECT/CT imaging.^12, 18, 23^ In addition to the spatial resolution of the SPECT/CT system and the size of the region of interest (ROI), several other factors—such as the shape of the ROI, the activity concentration ratio between the ROI and adjacent regions, and the specific SPECT/CT acquisition and reconstruction parameters—also influence the degree to which PVE impact the image quality^11^

To develop PVCs in SPECT/CT imaging, a phantom can be imaged with various inserts of known small volumes, each filled with a precise amount of activity.^7, 20^ From the quantified image, volume-dependent recovery coefficients can be computed, representing the fraction of recovered activity relative to the true activity levels in a ROI. These coefficients indicate the extent to which recovered activity in the ROI is affected by PVEs. The recovery coefficients can be fitted and extrapolated to construct a ‘recovery curve’, which in principle can be used to apply a PVC to a ROI of any volume.

In this systematic review on quantitative ^177^Lu SPECT/CT imaging, we present an overview on the accuracy of ^177^Lu activity quantifications in SPECT/CT imaging, an identification of methods for an improved accuracy when imaging small volumes subject to PVEs, and an assessment of the role for PVCs in clinical practice.

## II Methods

### Search Methodology

We followed the Preferred Reporting Items for Systematic reviews and Meta-Analyses (PRISMA) guidelines.^24^ A literature search was conducted using MEDLINE, EMBASE, and Web of Science until September 2024. Our search terms included *single-photon emission computed tomography, lutetium, quantification*, and *accuracy* (Supplementary Material Table S1, S2). There were no language or document type restrictions imposed on our search, but identified studies not available in English were later excluded.

### Study Selection Process and Eligibility Criteria

After the removal of duplicate records, abstracts were screened to evaluate if the study reports on the accuracy of ^177^Lu activity quantifications in SPECT/CT imaging. After screening an abstract, the study was assessed for eligibility for inclusion based on the following criteria:

- Reporting on absolute values of a quantitative parameter to assess the accuracy of ^177^Lu activity quantifications in SPECT/CT imaging which are based upon physical SPECT/CT measurements.
- Data reported from original study.

Full-text articles of potentially eligible studies were sought for retrieval and evaluated to ensure that all inclusion criteria were satisfied. Data from included studies was then extracted onto a standardised form (Supplementary Material Table S3).

## III Results

### Study Selection

The search strategy initially identified 616 records, of which 449 were unique, leaving 140 full-text articles to evaluate. In total, 46 studies met the inclusion criteria (Figure 1).

**Figure 1:**
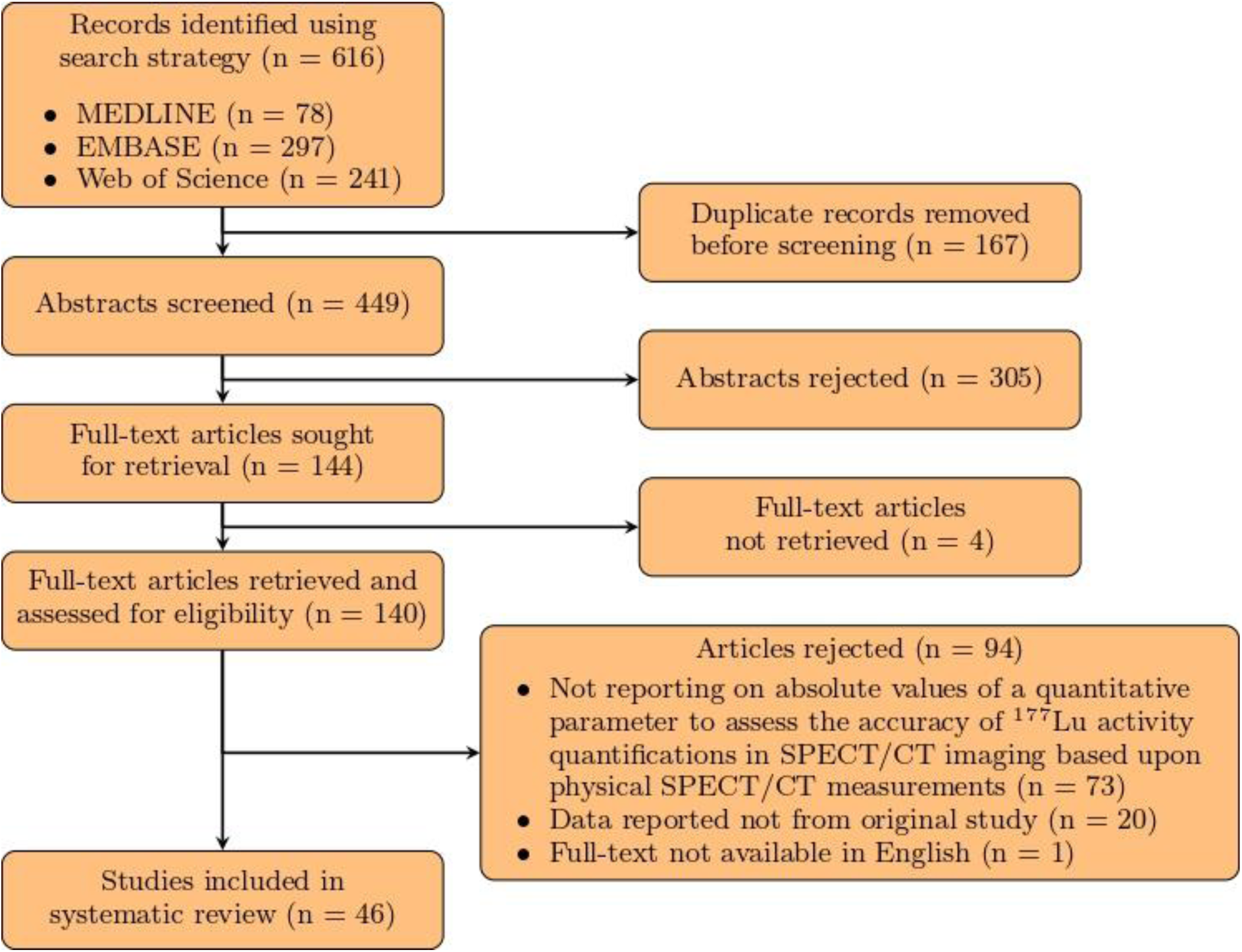
Flowchart showing the literature search strategy.

### Study Characteristics

Among the 46 included studies, distinct methods and materials have been adopted by authors to assess the accuracy of quantitative ^177^Lu SPECT/CT imaging. Firstly, studies can be characterised on the type of object imaged. Thirty-three studies ^6, 8, 9, 11, 16, 19, 21, 23, 25–49^ imaged common phantoms containing inserts with smooth geometric shapes such as cylinders or spheres. Eighteen studies ^6, 11–13, 15, 21, 22, 31, 36, 45, 46, 50–56^ imaged an anthropomorphic phantom composed of coarsely-edged, organ-shaped inserts for a better representation of clinical practice, which includes populations of patients with varying anatomical structures and internal distributions of ^177^Lu activity. To overcome all remaining inherent limits of mimicking clinical practice through phantoms, six studies ^19, 30, 40, 43, 57, 58^ imaged groups of patients.

Additionally, studies can be characterised on the quantitative parameters that have been reported. Thirty-one studies ^6, 8, 11, 12, 15, 16, 19, 21–23, 26, 27, 29–32, 34, 36, 39, 40, 42–46, 51–53, 55, 57, 58^ reported the percentage error of quantifications (i.e., deviations between SPECT/CT based and true ^177^Lu activity levels). Moreover, 25 studies ^6, 9, 11–13, 21, 23, 25, 28, 31, 33, 35, 37, 38, 40–42, 45, 47–50, 54, 56, 58^ reported recovery coefficients to evaluate the recovery of ^177^Lu activity when SPECT/CT imaging small volumes subject to PVEs. Seven studies ^21, 23, 34, 35, 38–40^ additionally reported coefficients of variation to assess the uniformity in SPECT/CT based ^177^Lu activity levels within a homogeneously filled ROI. As the data available on this parameter is limited, this parameter will not be discussed in the remainder of this review.

### Percentage Errors

Of the 31 studies that reported percentage errors of quantifications, there were 25 studies ^6, 11, 12, 15, 16, 19, 21–23, 26, 27, 30, 32, 34, 36, 39, 40, 42–46, 51, 52, 55, 58^ that allowed for the extraction of the minimum, average, and maximum value (Figure 2). There were six studies ^11, 16, 22, 30, 32, 44, 45^ in which the range of encountered percentage errors exceeded 100%.

**Figure 2:**
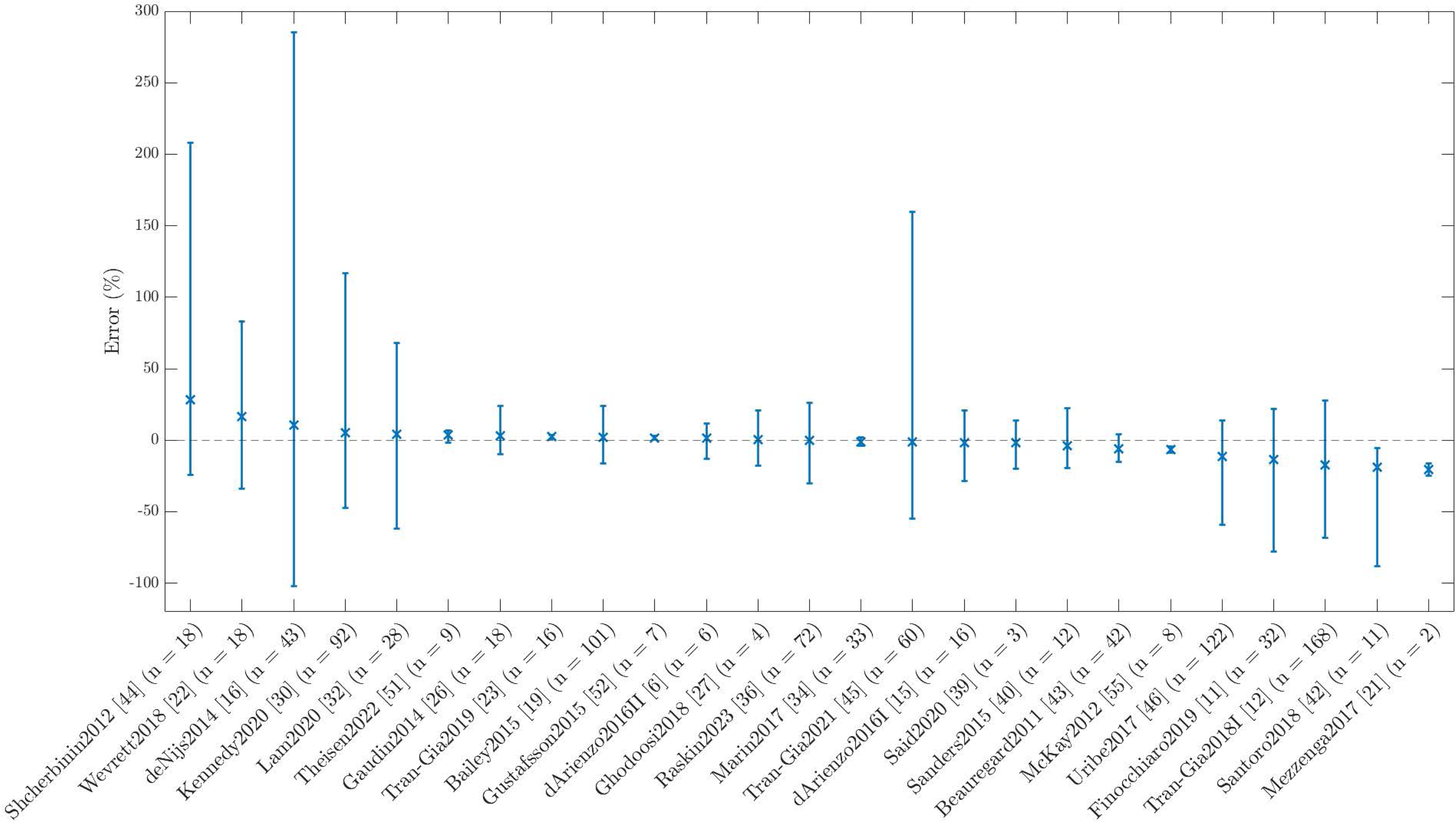
Minimum, average, and maximum percentage error of 177Lu activity quantifications in SPECT/CT imaging.

### Recovery Coefficients

Of the 25 studies that reported recovery coefficients, there were 20 studies ^6, 9, 11, 13, 21, 23, 25, 28, 33, 35, 38, 40–42, 45, 47–49, 54, 56^ that computed coefficients which can be directly compared from inserts with a common volume (i.e., volumes imaged in multiple studies on quantitative ^177^Lu SPECT/CT imaging). Figure 3 shows a full overview of minimum, average, and maximum recovery coefficients obtained in the collection of these studies, for volumes ranging from 0.5 to 26.5 mL. Specifications on the number of studies and measurements that are included for each volume can be found in the Supplementary Material (Table S4, S5). The general trend of an increasing impact of PVEs for a volume decreasing in size can clearly be observed with the average recovery coefficient gradually decreasing from 0.68 for a volume of 26.5 mL to 0.20 for a volume of 0.5 mL. A full recovery of ^177^Lu activity inserted in volumes smaller than 8 mL has not ever been achieved. Neither has the portion of ^177^Lu activity recovered from the smallest volume ever exceeded 50%.

**Figure 3:**
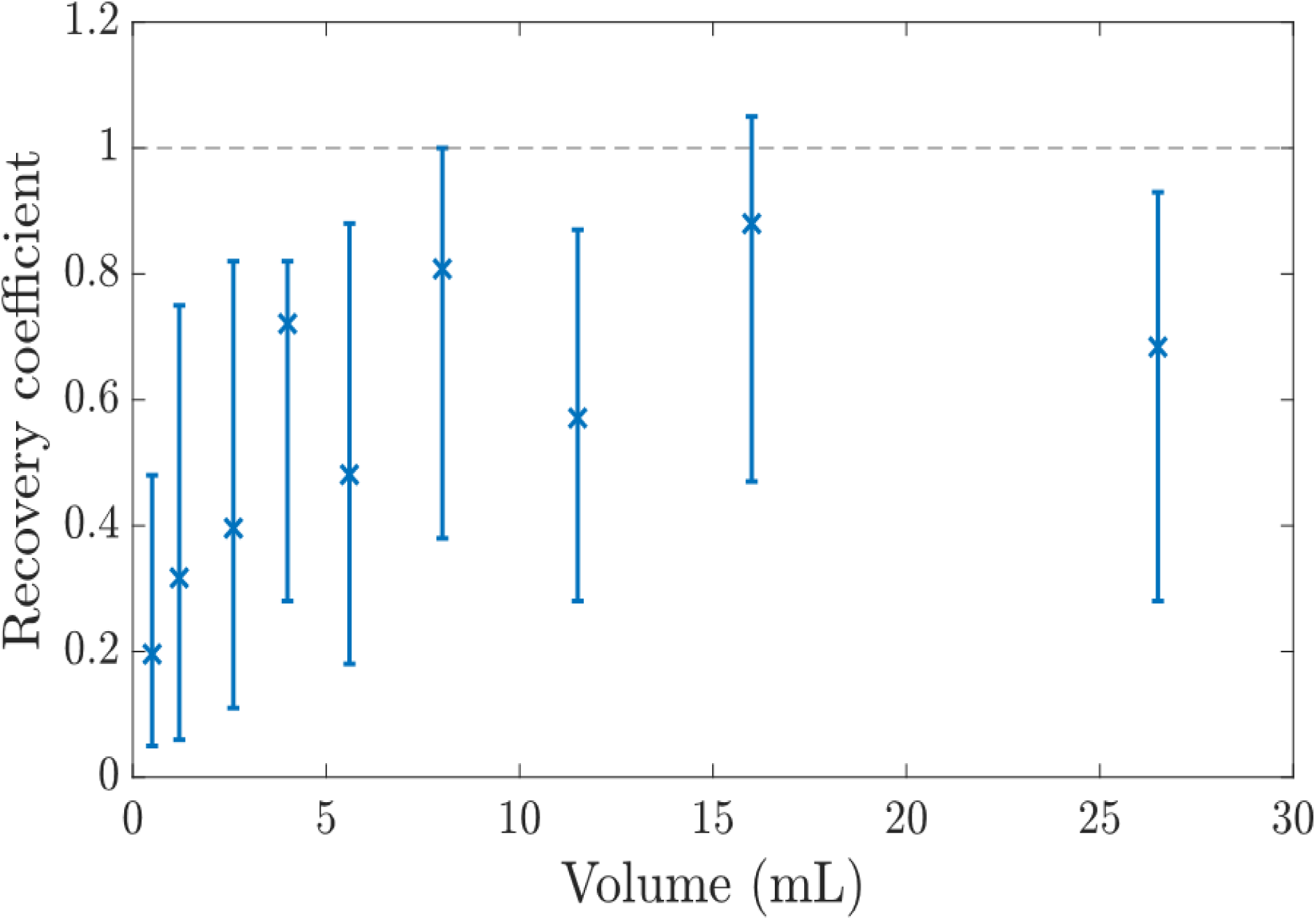
Minimum, average, and maximum recovery coefficients from common volumes in quantitative 177Lu SPECT/CT imaging.

### Methods for Improved Recovery Coefficients

A few methods for an improved recovery of ^177^Lu activity when SPECT/CT imaging small volumes subject to PVEs have been proposed in the literature. These include the implementation of correction methods ^28, 45, 54^ and an increased number of iterations and subsets [9, 21, 40] in the reconstruction process, the acquisition of primary emission counts around ^177^Lu’s gamma- emission at an energy of 208 keV only ^28, 40^, and imaging with a Cadmium Zinc Telluride (CZT)- based SPECT/CT system.^25, 41^

### Influence of Publication Date and Scanner Vendor on Recovery Coefficients

To characterise recent progress in the field of quantitative ^177^Lu SPECT/CT imaging, and the performance of SPECT/CT systems from distinct scanner vendors, subgroups have been taken from the data presented in Figure 3. In Figure 4, previously presented recovery coefficients are grouped by publication year and scanner vendor. Specifications on the number of studies and measurements that are included for each volume can be found in the Supplementary Material (Table S6-S9).

**Figure 4:**
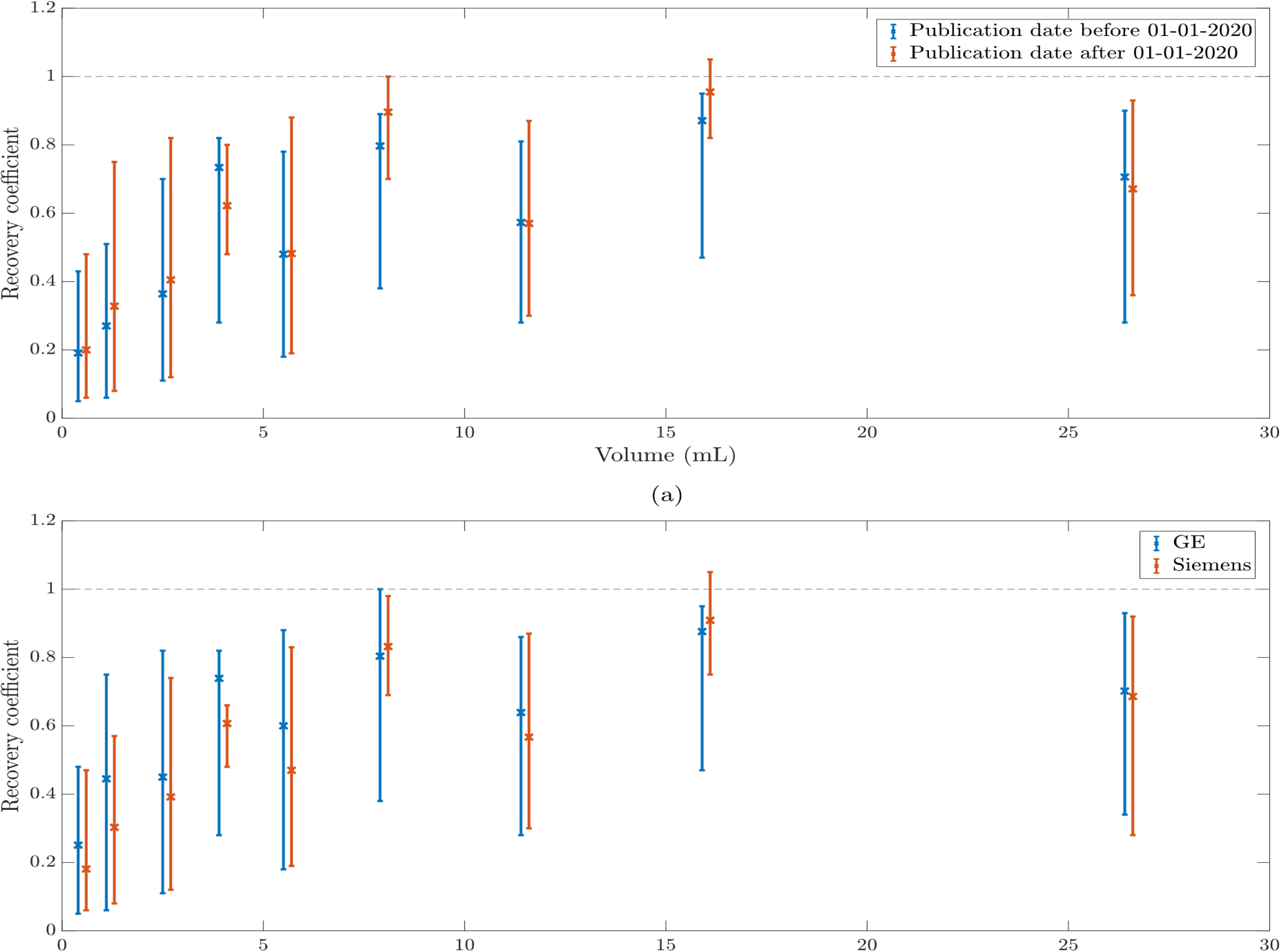
Recovery coefficients grouped by a) publication date, and b) scanner vendor.

In studies published after January 1, 2020, the maximum recovery coefficient was on average 13.3% larger than in those published before 2020 (Figure 4a). Performances of GE and Siemens SPECT/CT systems were compared as these were the only scanner vendors with data from multiple studies (Figure 4b). The maximum recovery coefficients obtained on a GE system were on average 7.5% larger than those from a Siemens system.

### Partial Volume Corrections

There were two studies ^11, 45^ that have made use of recovery coefficient measurements to study the influence of PVCs on the accuracy of quantitative ^177^Lu SPECT/CT imaging when imaging small volumes. After the construction of a recovery curve through phantom imaging, the influence of PVCs was investigated by imaging another phantom with differently sized inserts. The validation phantom was quantified twice; once with, and once without applying the previously established PVCs. Figure 5 shows the percentage error of quantifications, in a similar manner as was done in Figure 2, with and without the application of PVCs. To highlight the potential of PVCs, the data resulting from a measurement in Tran-Gia et al.^45^ where a PVC had been applied to an insert in the validation phantom which was evidently impacted differently by PVEs than the inserts used to establish the PVCs, has been excluded from the figure. In Figure 5, the accuracy, as well as the precision of ^177^Lu activity quantifications, was improved considerably by applying PVCs. Finocchiaro et al.^11^ realised a shift in the average percentage error from −28.4 to +1.5% while the total range of percentage errors decreased from 74 to 47%. Likewise, Tran-Gia et al. [45] realised a shift in the average percentage error from −31.2 to −14.3% and a decrease in percentage error range from 45 to 30%.

**Figure 5:**
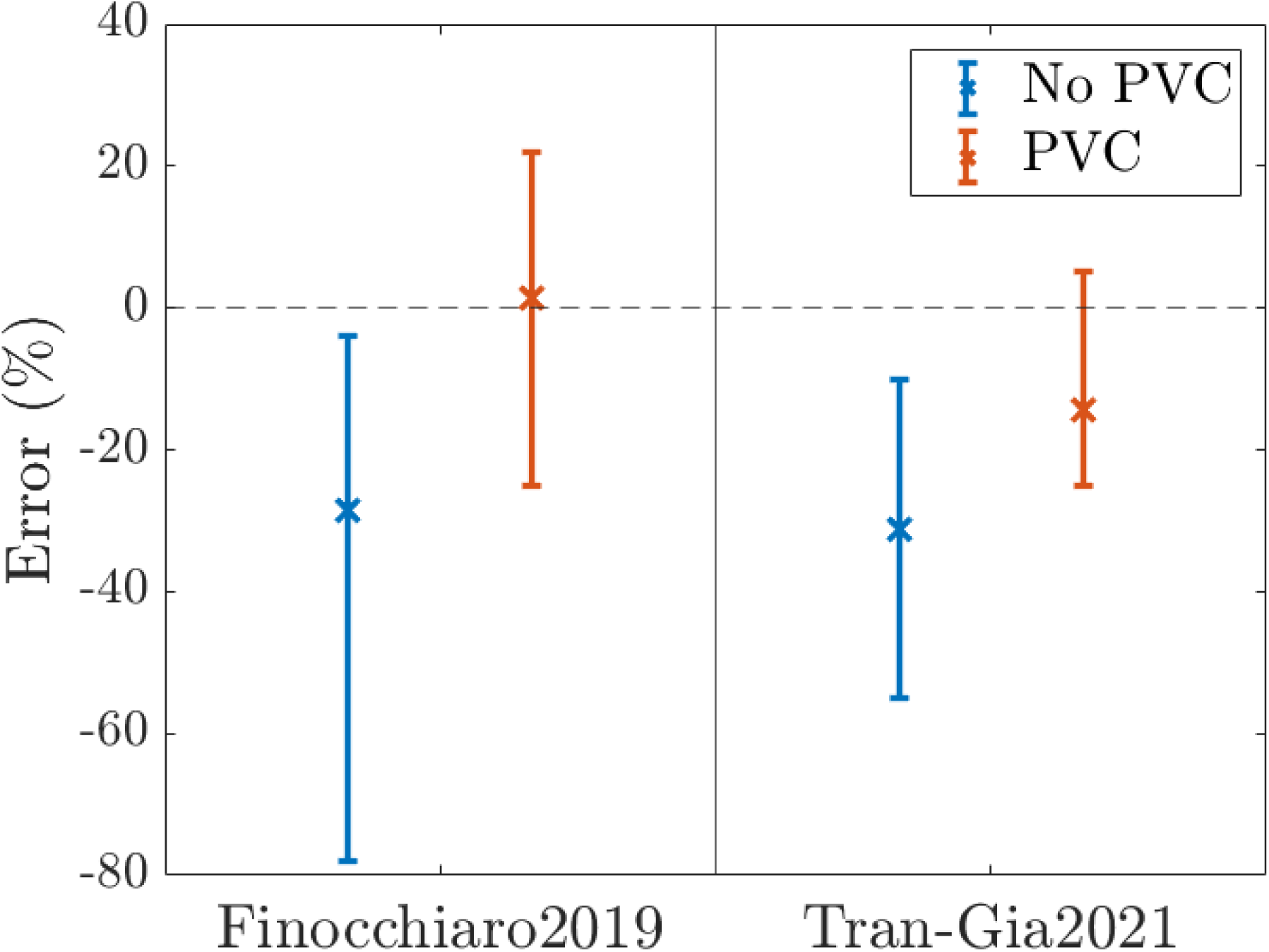
Minimum, average, and maximum percentage error of PVC-separated ^177^Lu activity quantifications in SPECT/CT imaging. Data was extracted from 16 measurements in Finocchiaro et al. [11], and 30 measurements in Tran-Gia et al. [45]. A partial exclusion of data meant that in Tran-Gia et al., partial-volume-corrected data was extracted from 25 measurements.

## IV Discussion

This is the first systematic review to thoroughly evaluate the accuracy of ^177^Lu activity quantifications in SPECT/CT imaging. A few reviews have briefly touched upon the accuracy of quantitative ^177^Lu SPECT/CT imaging before.^7, 14, 20^ However, the number of studies included in these reviews is limited. Besides presenting a full overview on the accuracy of quantitative ^177^Lu SPECT/CT imaging, this study summarises methods for an improved recovery of ^177^Lu activity when imaging small volumes subject to PVEs, and presented the influence of PVCs on the accuracy of quantitative ^177^Lu SPECT/CT imaging. Percentage errors of quantifications in studies on quantitative ^177^Lu SPECT/CT imaging were found to constitute a large range, repeatedly exceeding 100% within a single study. The recovery of ^177^Lu activity from small volumes was found to be limited, despite recent advances in quantitative SPECT/CT imaging and regardless of the scanner vendor. The application of PVCs was found to allow for improvements in the accuracy and precision of quantifications on small volumes.

In Figure 2, we observed that an accurate quantification of ^177^Lu activity in SPECT/CT imaging can be achieved both in phantom imaging as well as in patient images. However, it is important to note that the accuracy of quantification across studies should not be judged solely based on the figure. The range and average of percentage errors are highly dependent on the study design and its objectives. For example, studies focused on optimizing imaging procedures are expected to show a larger range and higher average of percentage errors compared to studies that utilize already optimized methods. Therefore, the accuracy of quantification in any given study must be interpreted in the context of its specific design and aims. Despite its potential, quantitative ^177^Lu SPECT/CT imaging is not yet widely established in clinical practice, partly because it can be a time- and cost-intensive process. Figure 2 showed that the lack of standardised protocols has resulted in large variations in the accuracy of quantifications due to the myriad of imaging hardware, calibration methods, acquisition protocols, and reconstruction algorithms used, which has obstructed the comparison of quantitative data from multi-centre studies. Although there is no specific set of imaging parameters which will lead to an optimal accuracy in all clinical applications, previous studies devoted to the optimisation of imaging parameters can provide a solid basis for a reduced variability in the accuracy of quantitative ^177^Lu SPECT/CT imaging through standardised procedures.^9, 45^

Figure 3 showed that the accuracy of quantitative ^177^Lu SPECT/CT images can be heavily degraded by PVEs, and thereby indicates the need for PVCs to allow for accurate quantitative ^177^Lu SPECT/CT imaging in clinical practice involving small lesions. The studies that were included in the figure can be divided in two groups. One group of studies imaged inserts with volumes of 0.5, 1.2, 2.6, 5.6, 11.5, and 26.5 mL, while the other group focused on inserts with volumes of 4.0, 8.0, and 16.0 mL. Due to the influence of multiple factors on PVE, the average recovery coefficient for a specific volume does not always increase with larger volumes. However, despite this variability, the general trend of an increasing recovery coefficient with larger volumes is clearly evident in the figure.

Literature has shown that the recovery of ^177^Lu activity can be greatly improved when implementing correction methods such as an attenuation correction, scatter correction, and resolution recovery, in the reconstruction process.^28, 45, 54^ Deviations between SPECT/CT-based and true activity levels arise from a combination of PVEs and other sources of error in the quantification process. These deviations can be significantly reduced by enhancing the accuracy of the reconstruction process. However, the application of these corrections may also introduce extreme under- or over-estimations of activity when incorrectly implemented.^16^ Furthermore, recovery coefficients can benefit from an increased number of iterations and subsets in the reconstruction process, especially for small volumes.^9, 21, 40^ However, the recovery will converge at some point, after which a further increased number of updates will only cause an increased build-up of noise, thereby degrading coefficients of variation.^21^ Hence, the optimal number of iterations and subsets is a compromise between the recovery of ^177^Lu activity and noise, and is application dependent. Additionally, acquiring counts from energy windows around the ^177^Lu gamma-emission energies of 113 and 208 keV naturally enhances the sensitivity of the SPECT/CT system compared to using a single energy window. However, collecting counts from both 113 keV and 208 keV windows simultaneously can degrade ^177^Lu activity recovery due to the introduction of additional noise.^28, 40^ A final method that was proposed in literature for an improved recovery of ^177^Lu activity from small volumes is to image with a CZT-based SPECT/CT system.^25, 41^ Unlike the previous methods, this approach involves modifying the imaging hardware by equipping the SPECT/CT system detectors with CZT scintillation crystals. These crystals are known for their superior energy resolution, although they come at a higher cost. Figure 4a demonstrates that recent advancements in quantitative 177Lu SPECT/CT imaging—such as the improved ability of reconstruction algorithms to incorporate the underlying physics of image formation, along with efforts to optimize imaging methods—have led to enhanced accuracy in quantifying small volumes. Future developments aimed at improving the spatial resolution of SPECT/CT systems may further enhance the recovery of 177Lu activity from small volumes. However, quantifications of the smallest volumes will inevitably be influenced by partial volume effects (PVEs) to some degree. As such, partial volume corrections (PVCs) will continue to play a key role in improving the accuracy of quantifications for small volumes, provided they are correctly implemented. The comparison of recovery coefficients between GE and Siemens SPECT/CT systems in Figure 4b showed no significant differences between the two imaging systemLastly, Figure 5 illustrates that substantial improvements in both the accuracy and precision of quantitative 177Lu SPECT/CT imaging can be achieved through the application of partial volume corrections (PVCs). Although these improvements have thus far been demonstrated in phantom imaging, they suggest that PVCs could play an important role in the clinical practice of quantitative 177Lu SPECT/CT imaging. While many studies have recognized that partial volume effects (PVEs) can significantly impact quantification accuracy, only two studies have explicitly investigated the effect of PVCs on this accuracy. As such, there is still insufficient evidence to conclusively establish the full impact of PVCs. Given that PVEs are influenced by various factors and the complexities of clinical practice, phantoms may not be able to fully capture all the nuances of PVEs across every clinical scenario. While anthropomorphic phantoms may provide a more realistic foundation for developing PVCs with potentially greater relevance to clinical practice compared to standard phantoms, the effectiveness of phantom-derived PVCs in actual clinical settings still requires further exploration.

### Strengths & Limitations

A strength of this systematic review is the comprehensive literature search strategy that was used to identify studies. Moreover, this is the first time that data from multiple studies on the accuracy of ^177^Lu activity quantifications when SPECT/CT imaging small volumes subject to PVEs has been compared. This review identifies pitfalls and measures to be overcome limitations.

A major limitation of this review is that all data included was acquired from single measurements, thereby limiting the reproducibility and statistical power of the findings. Moreover, keeping track of the propagation of uncertainties in measurements throughout the entire quantification process is currently not standard in the field of nuclear medicine ^22, 45^, which limits the ability to compare results from different studies. A final major limitation of this review is that PVCs have not been validated in patients yet, which is a prerequisite to fully assess the role for PVCs in quantitative ^177^Lu SPECT/CT imaging.

## V Conclusion

In conclusion, this systematic review found a large variation in the accuracy of ^177^Lu activity quantifications in SPECT/CT imaging. In particular, an inferior accuracy was observed when performing quantifications on small volumes subject to PVEs. The data suggests that a standardisation of quantitative imaging procedures and the application of PVCs may lead to an improved accuracy and precision of quantitative ^177^Lu SPECT/CT imaging, thereby allowing for the reliable determination of dose-response relationships for the further development of ^177^Lu- based PRRT.

## Statements & Declarations

## Author contributions

Julian van Oorschodt, Guy D. Eslick, and Enid Eslick contributed to the literature search, and study design. Carola van Pul, Martin Ugander and Enid Eslick supervised the process. Julian van Oorschodt performed the eligibility evaluation, data extraction, and data analysis. The first draft of the manuscript was written by Julian van Oorschodt, and all authors commented on previous versions of the manuscript. All authors read and approved the final manuscript.

## Data availability

The datasets generated during and/or analysed during the current study are available from the corresponding author on reasonable request.

## Ethics approval

Not applicable.

## Consent to participate

Not applicable.

## Consent for publication

Not applicable.

## Supplementary Material

**Table S1:** MEDLINE and EMBASE search strategy.

|  |  |
| --- | --- |
| 1 | "gamma camera" |
| 2 | SPECT |
| 3 | "single-photon emission computed tomography" |
| 4 | "single-photon emission computeri#ed tomography" |
| 5 | 1 or 2 or 3 or 4 |
| 6 | lutetium |
| 7 | lu |
| 8 | lu177 |
| 9 | 177lu |
| 10 | 6 or 7 or 8 or 9 |
| 11 | quantitative |
| 12 | quantification |
| 13 | quantifiable |
| 14 | quantitation |
| 15 | quanti#ed |
| 16 | 11 or 12 or 13 or 14 or 15 |
| 17 | accuracy |
| 18 | optimi#ation |
| 19 | optimi#ed |
| 20 | reconstruction |
| 21 | "recovery coefficient?" |
| 22 | 17 or 18 or 19 or 20 or 21 |
| 23 | 5 and 10 and 16 and 22 |

**Table S2:**
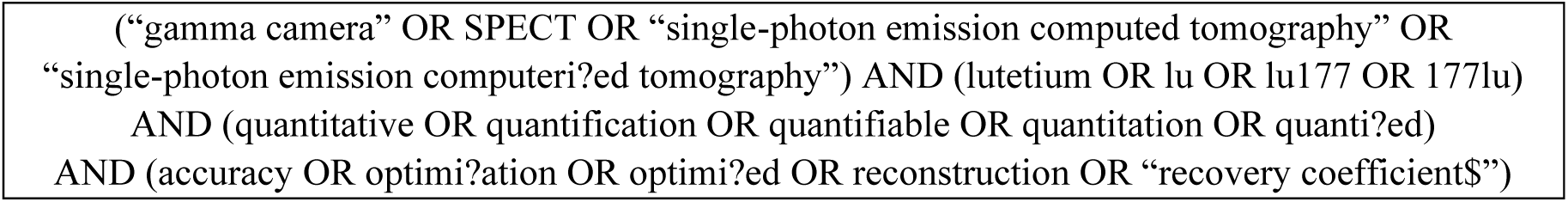
Web of Science search strategy.

**Table S3:** Overview of recorded data items.

|  |  |  |
| --- | --- | --- |
| **General Items**<br>• Author name<br>• Year of publication<br>• Country/continent | **Measurement Hardware**<br>• SPECT/CT scanner vendor/model<br>• Dose calibrator vendor/model<br>• Traceability activity | **CT Acquisition**<br>• Tube current<br>• Tube voltage<br>• Slice thickness |
| **SPECT Acquisition**<br>• Collimator<br>• Energy windows<br>• Matrix size<br>• Pixel size<br>• Number of detectors<br>• Detector orbit<br>• Acquisition mode<br>• Number of views<br>• Time per view<br>• Sampling angle<br>• Total rotation angle | **SPECT/CT Reconstruction**<br>• Reconstruction software<br>• Underlying algorithm<br>• Attenuation correction<br>• Scatter correction<br>• Resolution recovery<br>• Other corrections<br>• Number of iterations<br>• Number of subsets<br>• Filter<br>• ROI drawing |  |
| **Imaging Objects**<br>• Object to derive conversion factor from<br>• Object to evaluate accuracy from<br>• Inserted activity level/concentration/ratio | **Accuracy Results**<br>• Percentage errors<br>• Recovery coefficients<br>• Coefficients of variation |  |

**Table S4:**
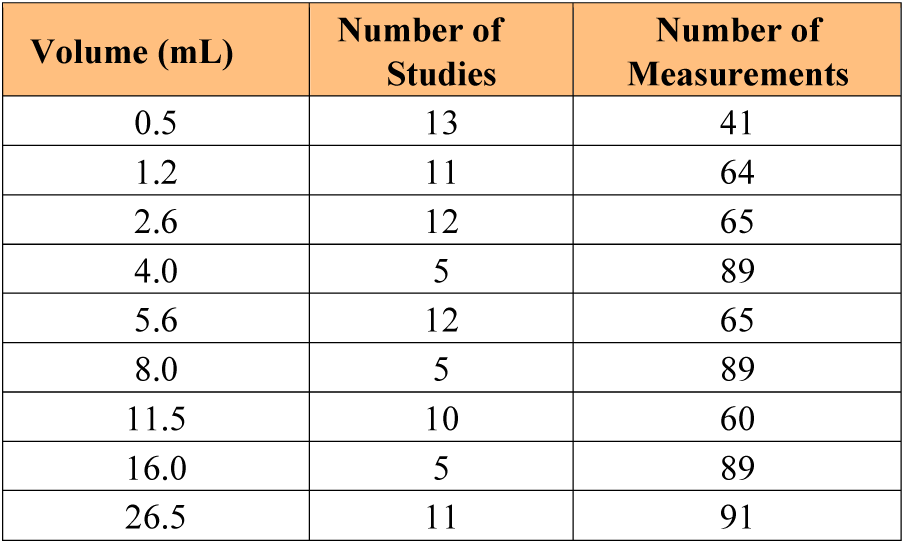
Overview of the numbers of studies and measurements contributing to data in Figure 3.

| Volume (mL) | Number of Studies | Number of Measurements |
| --- | --- | --- |
| 0.5 | 13 | 41 |
| 1.2 | 11 | 64 |
| 2.6 | 12 | 65 |
| 4.0 | 5 | 89 |
| 5.6 | 12 | 65 |
| 8.0 | 5 | 89 |
| 11.5 | 10 | 60 |
| 16.0 | 5 | 89 |
| 26.5 | 11 | 91 |

**Table S5:** Overview of the numbers of measurements per study contributing to data in Figure 3.

| V = 0.5 mL |  | V = 1.2 mL |  | V = 2.6 mL |  |
| --- | --- | --- | --- | --- | --- |
| Study | Number of Measurements | Study | Number of Measurements | Study | Number of Measurements |
| dArienzo2016II<sup>[6]</sup> | 1 | dArienzo2016II<sup>[6]</sup> | 1 | dAmbrosio2013<sup>[47]</sup> | 2 |
| Delker2016<sup>[48]</sup> | 1 | Delker2016<sup>[48]</sup> | 1 | dArienzo2016II<sup>[6]</sup> | 1 |
| Ferrando2021<sup>[49]</sup> | 1 | Hippelainen2016<sup>[54]</sup> | 3 | Delker2016<sup>[48]</sup> | 1 |
| Finnochiaro2019<sup>[11]</sup> | 1 | Huizing2020<sup>[28]</sup> | 36 | Hippelainen2016<sup>[54]</sup> | 3 |
| Leube2022<sup>[33]</sup> | 1 | Leube2022<sup>[33]</sup> | 1 | Huizing2020<sup>[28]</sup> | 36 |
| Marquis2021<sup>[35]</sup> | 3 | Marquis2021<sup>[35]</sup> | 3 | Leube2022<sup>[33]</sup> | 1 |
| Peters2020<sup>[9]</sup> | 10 | Merrick2021<sup>[56]</sup> | 2 | Marquis2021<sup>[35]</sup> | 3 |
| Ryu2019<sup>[38]</sup> | 1 | Ryu2019<sup>[38]</sup> | 1 | Merrick2021<sup>[56]</sup> | 1 |
| Sanders2015<sup>[40]</sup> | 4 | Sandstrom2019<sup>[41]</sup> | 2 | Ryu2019<sup>[38]</sup> | 1 |
| Sandstrom2019<sup>[41]</sup> | 2 | Tran-Gia2019<sup>[23]</sup> | 4 | Sandstrom2019<sup>[41]</sup> | 2 |
| Santoro2018<sup>[42]</sup> | 2 | Tran-Gia2021<sup>[45]</sup> | 10 | Tran-Gia2019<sup>[23]</sup> | 4 |
| Tran-Gia2019<sup>[23]</sup> | 4 |  |  | Tran-Gia2021<sup>[45]</sup> | 10 |
| Tran-Gia2021<sup>[45]</sup> | 10 |  |  |  |  |

| V = 4.0 mL |  | V = 5.6 mL |  | V = 8.0 mL |  |
| --- | --- | --- | --- | --- | --- |
| Study | Number of Measurements | Study | Number of Measurements | Study | Number of Measurements |
| Mezzenga2017<sup>[21]</sup> | 72 | dArienzo2016II<sup>[6]</sup> | 1 | Mezzenga2017<sup>[21]</sup> | 72 |
| Peters2020<sup>[9]</sup> | 10 | Delker2016<sup>[48]</sup> | 1 | Peters2020<sup>[9]</sup> | 10 |
| Robinson2016II<sup>[13]</sup> | 1 | Finnochiaro2019<sup>[11]</sup> | 1 | Robinson2016II<sup>[13]</sup> | 1 |
| Sanders2015<sup>[40]</sup> | 4 | Hippelainen2016<sup>[54]</sup> | 3 | Sanders2015<sup>[40]</sup> | 4 |
| Santoro2018<sup>[42]</sup> | 2 | Huizing2020<sup>[28]</sup> | 36 | Santoro2018<sup>[42]</sup> | 2 |
|  |  | Leube2022<sup>[33]</sup> | 1 |  |  |
|  |  | Marquis2021<sup>[35]</sup> | 3 |  |  |
|  |  | Merrick2021<sup>[56]</sup> | 2 |  |  |
|  |  | Ryu2019<sup>[38]</sup> | 1 |  |  |
|  |  | Sandstrom2019<sup>[41]</sup> | 2 |  |  |
|  |  | Tran-Gia2019<sup>[23]</sup> | 4 |  |  |
|  |  | Tran-Gia2021<sup>[45]</sup> | 10 |  |  |

| V = 11.5 mL |  | V = 16.0 mL |  | V = 26.5 mL |  |
| --- | --- | --- | --- | --- | --- |
| Study | Number of Measurements | Study | Number of Measurements | Study | Number of Measurements |
| dArienzo2016II<sup>[6]</sup> | 1 | Mezzenga2017<sup>[21]</sup> | 72 | Badel2022<sup>[25]</sup> | 6 |
| Delker2016<sup>[48]</sup> | 1 | Peters2020<sup>[9]</sup> | 10 | dAmbrosio2013<sup>[47]</sup> | 2 |
| Huizing2020<sup>[28]</sup> | 36 | Robinson2016II<sup>[13]</sup> | 1 | dArienzo2016II<sup>[6]</sup> | 1 |
| Leube2022<sup>[33]</sup> | 1 | Sanders2015<sup>[40]</sup> | 4 | Delker2016<sup>[48]</sup> | 1 |
| Marquis2021<sup>[35]</sup> | 3 | Santoro2018<sup>[42]</sup> | 2 | Huizing2020<sup>[28]</sup> | 36 |
| Merrick2021<sup>[56]</sup> | 1 |  |  | Leube2022<sup>[33]</sup> | 1 |
| Ryu2019<sup>[38]</sup> | 1 |  |  | Marquis2021<sup>[35]</sup> | 3 |
| Sandstrom2019<sup>[41]</sup> | 2 |  |  | Merrick2021<sup>[56]</sup> | 1 |
| Tran-Gia2019<sup>[23]</sup> | 4 |  |  | Sandstrom2019<sup>[41]</sup> | 2 |
| Tran-Gia2021<sup>[45]</sup> | 10 |  |  | Tran-Gia2019<sup>[23]</sup> | 28 |
|  |  |  |  | Tran-Gia2021<sup>[45]</sup> | 10 |

**Table S6:**
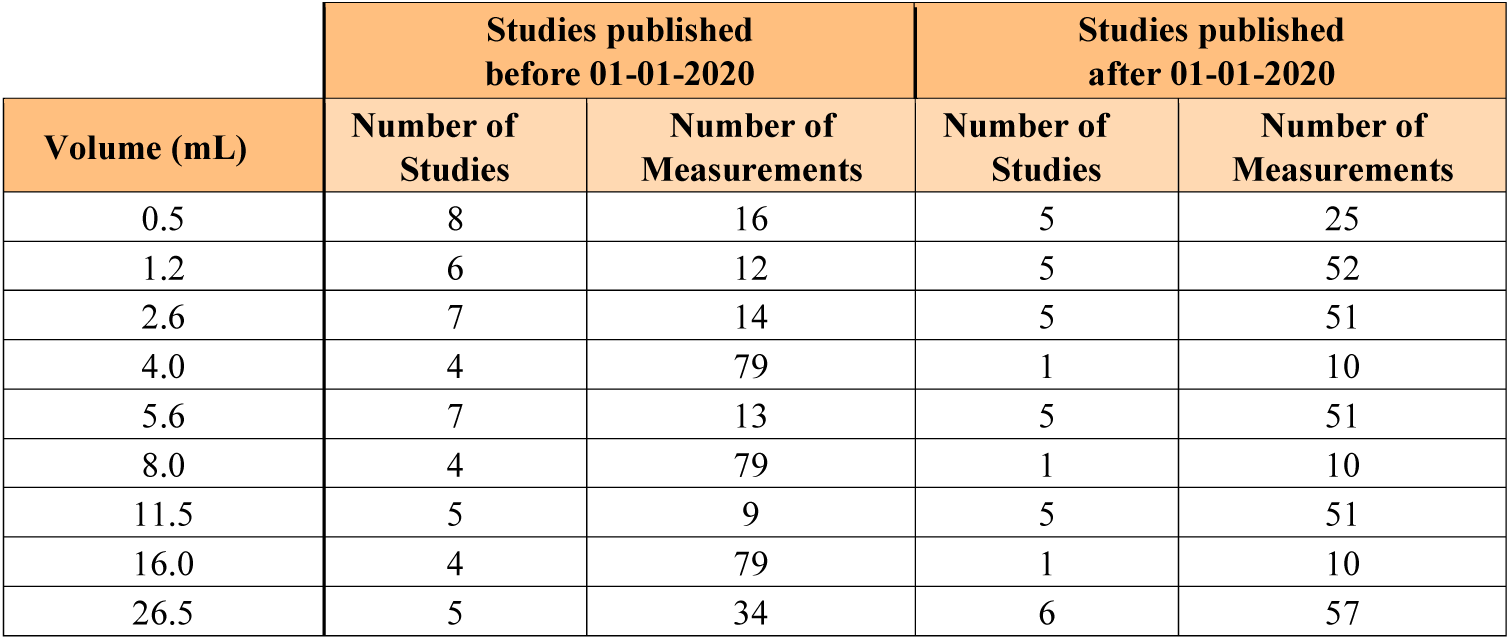
Overview of the numbers of studies and measurements contributing to data in Figure 4a.

| Volume (mL) | Studies published before 01-01-2020 |  | Studies published after 01-01-2020 |  |
| --- | --- | --- | --- | --- |
|  | Number of Studies | Number of Measurements | Number of Studies | Number of Measurements |
| 0.5 | 8 | 16 | 5 | 25 |
| 1.2 | 6 | 12 | 5 | 52 |
| 2.6 | 7 | 14 | 5 | 51 |
| 4.0 | 4 | 79 | 1 | 10 |
| 5.6 | 7 | 13 | 5 | 51 |
| 8.0 | 4 | 79 | 1 | 10 |
| 11.5 | 5 | 9 | 5 | 51 |
| 16.0 | 4 | 79 | 1 | 10 |
| 26.5 | 5 | 34 | 6 | 57 |

**Table S7:**
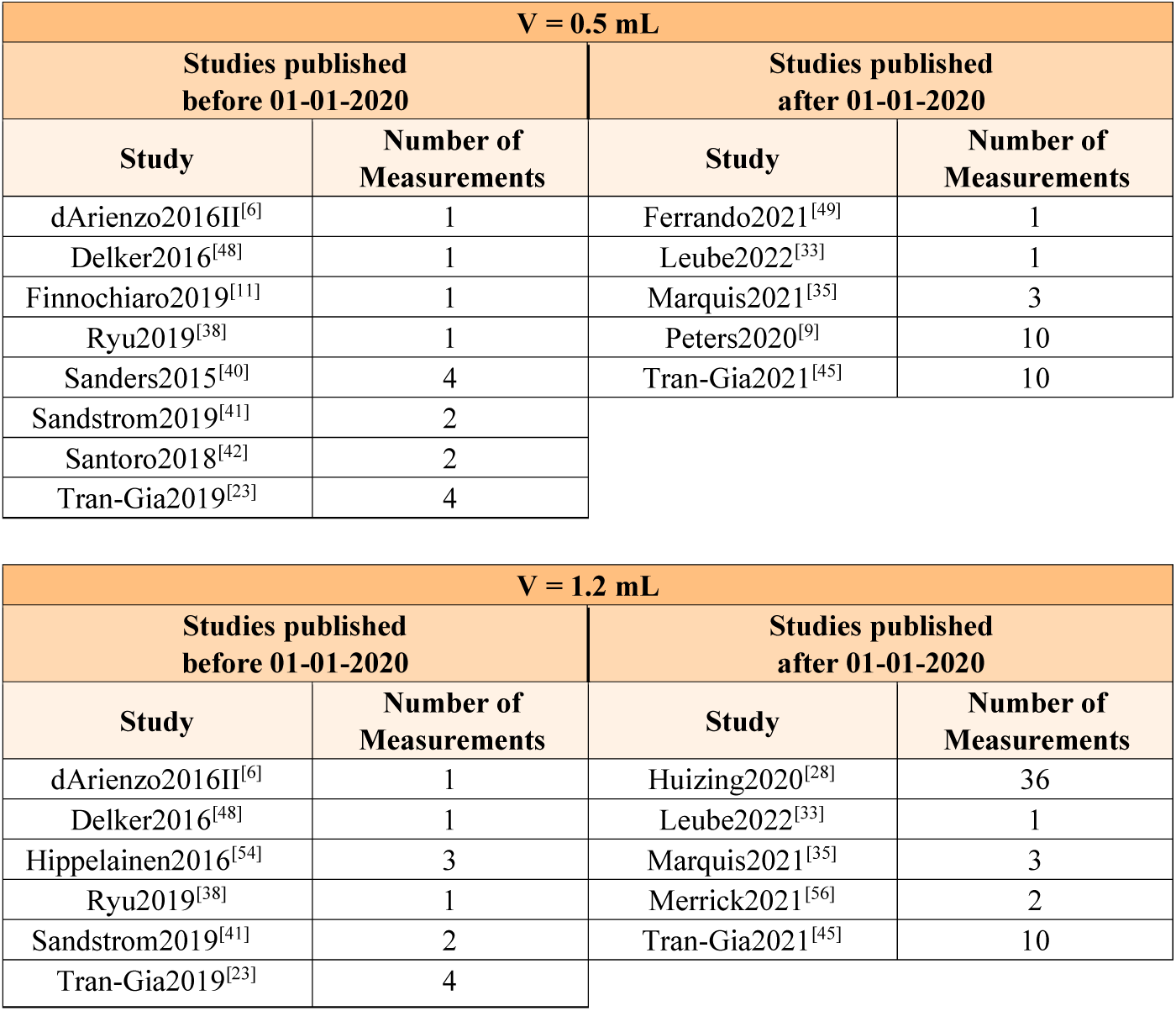
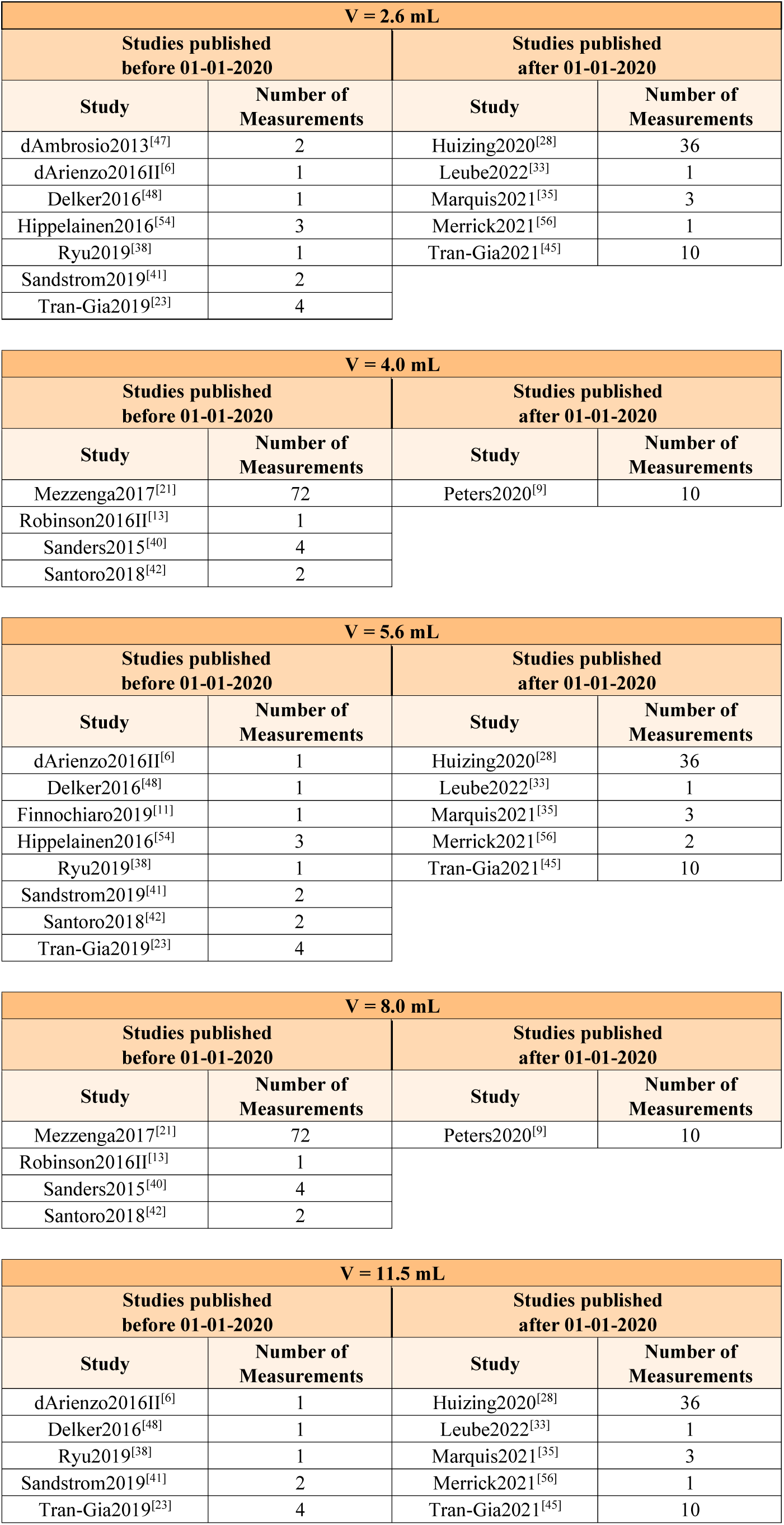
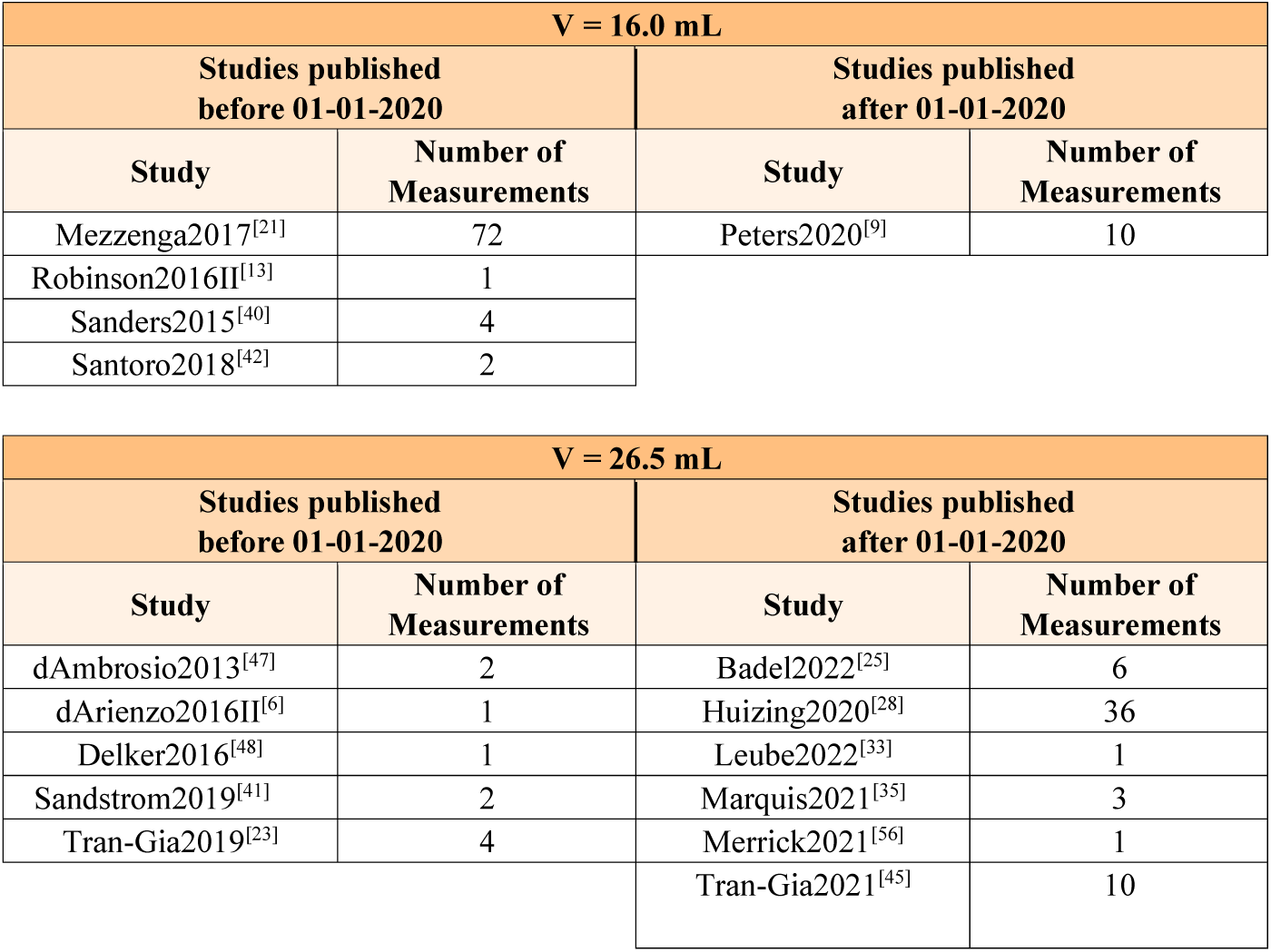
Overview of the numbers of measurements per study contributing to data in Figure 4a.

**Table S8:** Overview of the numbers of studies and measurements contributing to data in Figure 4b.

| Volume (mL) | Measurements on GE SPECT/CT system |  | Measurements on Siemens SPECT/CT system |  |
| --- | --- | --- | --- | --- |
|  | Number of Studies | Number of Measurements | Number of Studies | Number of Measurements |
| 0.5 | 4 | 12 | 10 | 27 |
| 1.2 | 2 | 8 | 9 | 54 |
| 2.6 | 3 | 10 | 9 | 53 |
| 4.0 | 4 | 77 | 2 | 12 |
| 5.6 | 2 | 8 | 10 | 55 |
| 8.0 | 4 | 77 | 2 | 12 |
| 11.5 | 2 | 8 | 8 | 50 |
| 16.0 | 4 | 77 | 2 | 12 |
| 26.5 | 3 | 10 | 7 | 73 |

**Table S9:** Overview of the numbers of measurements per study contributing to data in Figure 4b.

| V = 0.5 mL |  |  |  |
| --- | --- | --- | --- |
| Studies published before 01-01-2020 |  | Studies published after 01-01-2020 |  |
| Study | Number of Measurements | Study | Number of Measurements |
| Peters2020<sup>[9]</sup> | 2 | Delker2016<sup>[48]</sup> | 1 |
| Sandstrom2019<sup>[41]</sup> | 2 | Ferrando2021<sup>[49]</sup> | 1 |
| Santoro2018<sup>[42]</sup> | 2 | Finocchiario2019<sup>[11]</sup> | 1 |
| Tran-Gia2021<sup>[45]</sup> | 6 | Leube2022<sup>[33]</sup> | 1 |
|  |  | Marquis2021<sup>[35]</sup> | 3 |
|  |  | Peters2020<sup>[9]</sup> | 8 |
|  |  | Ryu2019<sup>[38]</sup> | 1 |
|  |  | Sanders2015<sup>[40]</sup> | 4 |
|  |  | Tran-Gia2019<sup>[23]</sup> | 4 |
|  |  | Tran-Gia2021<sup>[45]</sup> | 3 |

| V = 1.2 mL |  |  |  |
| --- | --- | --- | --- |
| Studies published before 01-01-2020 |  | Studies published after 01-01-2020 |  |
| Study | Number of Measurements | Study | Number of Measurements |
| Sandstrom2019<sup>[41]</sup> | 2 | Delker2016<sup>[48]</sup> | 1 |
| Tran-Gia2021<sup>[45]</sup> | 6 | Hippelainen2016<sup>[54]</sup> | 3 |
|  |  | Huizing2020<sup>[28]</sup> | 36 |
|  |  | Leube2022<sup>[33]</sup> | 1 |
|  |  | Marquis2021<sup>[35]</sup> | 3 |
|  |  | Merrick2021<sup>[56]</sup> | 2 |
|  |  | Ryu2019<sup>[38]</sup> | 1 |
|  |  | Tran-Gia2019<sup>[23]</sup> | 4 |
|  |  | Tran-Gia2021<sup>[45]</sup> | 3 |

| V = 2.6 mL |  |  |  |
| --- | --- | --- | --- |
| Studies published before 01-01-2020 |  | Studies published after 01-01-2020 |  |
| Study | Number of Measurements | Study | Number of Measurements |
| dAmbrosio2013<sup>[47]</sup> | 2 | Delker2016<sup>[48]</sup> | 1 |
| Sandstrom2019<sup>[41]</sup> | 2 | Hippelainen2016<sup>[54]</sup> | 3 |
| Tran-Gia2021<sup>[45]</sup> | 6 | Huizing2020<sup>[28]</sup> | 36 |
|  |  | Leube2022<sup>[33]</sup> | 1 |
|  |  | Marquis2021<sup>[35]</sup> | 3 |
|  |  | Merrick2021<sup>[56]</sup> | 1 |
|  |  | Ryu2019<sup>[38]</sup> | 1 |
|  |  | Tran-Gia2019<sup>[23]</sup> | 4 |
|  |  | Tran-Gia2021<sup>[45]</sup> | 3 |

| V = 4.0 mL |  |  |  |
| --- | --- | --- | --- |
| Studies published before 01-01-2020 |  | Studies published after 01-01-2020 |  |
| Study | Number of Measurements | Study | Number of Measurements |
| Mezzenga2017<sup>[21]</sup> | 72 | Peters2020<sup>[9]</sup> | 8 |
| Peters2020<sup>[9]</sup> | 2 | Sanders2015<sup>[40]</sup> | 4 |
| Robinson2016II<sup>[13]</sup> | 1 |  |  |
| Santoro2018<sup>[42]</sup> | 2 |  |  |

| V = 5.6 mL |  |  |  |
| --- | --- | --- | --- |
| Studies published before 01-01-2020 |  | Studies published after 01-01-2020 |  |
| Study | Number of Measurements | Study | Number of Measurements |
| Sandstrom2019<sup>[41]</sup> | 2 | Delker2016<sup>[48]</sup> | 1 |
| Tran-Gia2021<sup>[45]</sup> | 6 | Finocchiario2019<sup>[11]</sup> | 1 |
|  |  | Hippelainen2016<sup>[54]</sup> | 3 |
|  |  | Huizing2020<sup>[28]</sup> | 36 |
|  |  | Leube2022<sup>[33]</sup> | 1 |
|  |  | Marquis2021<sup>[35]</sup> | 3 |
|  |  | Merrick2021<sup>[56]</sup> | 2 |
|  |  | Ryu2019<sup>[38]</sup> | 1 |
|  |  | Tran-Gia2019<sup>[23]</sup> | 4 |
|  |  | Tran-Gia2021<sup>[45]</sup> | 3 |

| V = 8.0 mL |  |  |  |
| --- | --- | --- | --- |
| Studies published before 01-01-2020 |  | Studies published after 01-01-2020 |  |
| Study | Number of Measurements | Study | Number of Measurements |
| Mezzenga2017<sup>[21]</sup> | 72 | Peters2020<sup>[9]</sup> | 8 |
| Peters2020<sup>[9]</sup> | 2 | Sanders2015<sup>[40]</sup> | 4 |
| Robinson2016II<sup>[13]</sup> | 1 |  |  |
| Santoro2018<sup>[42]</sup> | 2 |  |  |

| V = 11.5 mL |  |  |  |
| --- | --- | --- | --- |
| Studies published before 01-01-2020 |  | Studies published after 01-01-2020 |  |
| Study | Number of Measurements | Study | Number of Measurements |
| Sandstrom2019<sup>[41]</sup> | 2 | Delker2016<sup>[48]</sup> | 1 |
| Tran-Gia2021<sup>[45]</sup> | 6 | Huizing2020<sup>[28]</sup> | 36 |
|  |  | Leube2022<sup>[33]</sup> | 1 |
|  |  | Marquis2021<sup>[35]</sup> | 3 |
|  |  | Merrick2021<sup>[56]</sup> | 1 |
|  |  | Ryu2019<sup>[38]</sup> | 1 |
|  |  | Tran-Gia2019<sup>[23]</sup> | 4 |
|  |  | Tran-Gia2021<sup>[45]</sup> | 3 |

| V = 16.0 mL |  |  |  |
| --- | --- | --- | --- |
| Studies published before 01-01-2020 |  | Studies published after 01-01-2020 |  |
| Study | Number of Measurements | Study | Number of Measurements |
| Mezzenga2017<sup>[21]</sup> | 72 | Peters2020<sup>[9]</sup> | 8 |
| Peters2020<sup>[9]</sup> | 2 | Sanders2015<sup>[40]</sup> | 4 |
| Robinson2016II<sup>[13]</sup> | 1 |  |  |
| Santoro2018<sup>[42]</sup> | 2 |  |  |

| V = 26.5 mL |  |  |  |
| --- | --- | --- | --- |
| Studies published before 01-01-2020 |  | Studies published after 01-01-2020 |  |
| Study | Number of Measurements | Study | Number of Measurements |
| dAmbrosio2013<sup>[47]</sup> | 2 | Delker2016<sup>[48]</sup> | 1 |
| Sandstrom2019<sup>[41]</sup> | 2 | Huizing2020<sup>[28]</sup> | 36 |
| Tran-Gia2021<sup>[45]</sup> | 6 | Leube2022<sup>[33]</sup> | 1 |
|  |  | Marquis2021<sup>[35]</sup> | 3 |
|  |  | Merrick2021<sup>[56]</sup> | 1 |
|  |  | Tran-Gia2019<sup>[23]</sup> | 28 |
|  |  | Tran-Gia2021<sup>[45]</sup> | 3 |

